# OnSIDES (ON-label SIDE effectS resource) Database : Extracting Adverse Drug Events from Drug Labels using Natural Language Processing Models

**DOI:** 10.1101/2024.03.22.24304724

**Authors:** Yutaro Tanaka, Hsin Yi Chen, Pietro Belloni, Undina Gisladottir, Jenna Kefeli, Jason Patterson, Apoorva Srinivasan, Michael Zietz, Gaurav Sirdeshmukh, Jacob Berkowitz, Kathleen LaRow Brown, Nicholas P. Tatonetti

## Abstract

Adverse drug events (ADEs) are the fourth leading cause of death in the US and cost billions of dollars annually in increased healthcare costs. However, few machine-readable databases of ADEs exist, limiting the opportunity to study drug safety on a broader, systematic scale. Recent advances in Natural Language Processing methods, such as BERT models, present an opportunity to accurately extract relevant information from unstructured biomedical text. As such, we fine-tuned a PubMedBERT model to extract ADE terms from descriptive text in FDA Structured Product Labels for prescription drugs. With this model, we achieve an F1 score of 0.90, AUROC of 0.92, and AUPR of 0.95 at extracting ADEs from the labels’ “Adverse Reactions”. We further utilize this method to extract serious ADEs from labels’ “Boxed Warnings”, and ADEs specifically noted for pediatric patients. Here, we present OnSIDES (ON-label SIDE effectS resource), a compiled, computable database of drug-ADE pairs generated with this method. OnSIDES contains more than 3.6 million drug-ADE pairs for 3,233 unique drug ingredient combinations extracted from 47,211 labels. Additionally, we expand this method to extract ADEs from drug labels of other major nations/regions - Japan, the UK, and the EU - to build a complementary OnSIDES-INTL database. To present potential applications, we used OnSIDES to predict novel drug targets and indications, analyze enrichment of ADEs across drug classes, and predict novel ADEs from chemical compound structures. We conclude that OnSIDES can be utilized as a comprehensive resource to study and enhance drug safety.

**One Sentence Summary:** OnSIDES is a large, comprehensive database of adverse drug events extracted from drug labels using natural language processing methods.

## Introduction

### Adverse Drug Events (ADEs)

Adverse Drug Events (ADEs) are broadly defined as unintended, harmful events related to the usage of medication (1), and are the fifth leading cause of death internationally (2). ADEs are a cause of significant avoidable additional healthcare costs, with approximately ½ of ADEs known to be preventable, and the annual cost of prescription drug-related morbidity and mortality in the US was estimated to be $528.4 billion in 2016 alone (3–4). While many drug safety studies and clinical trials have been conducted on specific medications, far fewer studies have studied the occurrence of ADEs more broadly, due to the heterogeneity and complexity of the phenomenon itself, and the lack of standardized data available (5–6). The comprehensive study of ADEs requires high-quality, machine-readable databases encompassing the breadth of drugs currently used in healthcare.

### ADE Databases

There are a number of ADE databases currently available for public usage, which are constructed by extracting, stratifying or detecting signals from a variety of sources, including drug labels, electronic health records, academic literature, social media, and adverse event reporting systems (7–11). In particular, drug labels remain the gold standard primary resource for ADE information that both physicians and patients reference, with many international regulatory agencies such as the US FDA (Food and Drug Administration) and the EMA (European Medicines Agency) maintaining publicly accessible repositories of drug labels (12). However, these drug label repositories are not necessarily standardized or optimized for machine readability, but rather a collection of individual files constituting the raw or formatted drug label text. Furthermore, as they are not limited to ADEs, containing all mandated drug information, it is not readily usable for the analysis of ADEs. One of the most widely utilized ADE databases derived from drug label data is SIDER, most recently updated in 2015 (13). It was constructed using a dictionary-based named entity recognition (NER) method to extract the adverse event terms from FDA Structured Product Labeling (SPLs) drug labels. However, no currently up-to-date, computable databases exist for on-label ADEs.

### Deep Learning Methods for Unstructured Biomedical Data

In recent years, significant advancements have been made in NLP (Natural Language Processing) methods, including the introduction and development of BERT (Bidirectional Encoder Representations from Transformers) models (14). By leveraging transformer architectures to learn text representations and contextual dependencies, BERT models have greatly improved performance in conducting language tasks such as sentiment analysis, keyword classification, and machine translation. In a biomedical context, numerous BERT models, such as BioBERT, MedBERT, and BiomedBERT, have been developed by training models on different relevant text corpora, such as EHR data and biomedical academic literature. (15–17). These models have been used for different tasks, such as biomedical named entity recognition, relations/association extraction, and answering questions. There have also been prior efforts to specifically leverage these methodological advancements in studying ADEs. Of note, the “Adverse Drug Reaction Extraction from Drug Labels” task of the Text Analysis Conference (TAC) 2017 was presented to encourage the development of computational methods to recognize mentions of ADEs from drug labels and their relationships. As part of this task, the organizers published a manually annotated, gold-standard reference dataset for extracting adverse drug events from 200 FDA SPLs (18–19). Building upon the TAC2017 task, a number of deep learning and NLP methods have been developed to extract ADE terms from drug label text (20–21). While these research efforts have presented promising results of high accuracy in extracting true ADE mentions, indicating the possibility of using NLP methods for this task, further challenges lie in leveraging these methods to systematically construct a comprehensive and readily accessible database.

### OnSIDES Database for ADEs

Leveraging advancements in NLP term extraction methods and building upon the methods proposed for the TAC2017 task, we fine-tuned a PubMedBERT, a BERT model pre-trained on biomedical research article abstracts and full-text articles from PubMed, to predict whether a given term in drug label text is an ADE related term (22). To do so, we first extract candidate ADE-related terms from the drug label using standard MedDRA vocabulary terms as a reference dictionary and then use the PubMedBERT model trained on a manually annotated set of drug labels to predict whether the possible term is an ADE for the given drug. We applied this method to all publicly available FDA SPLs to construct the OnSIDES (ON label SIDE effectS) database of both adverse reactions and boxed warnings. We plan for this database to be updated quarterly for SPL updates to incorporate all updated drug labels and new drugs to reflect all of the medications currently available. Furthermore, we adapted this method to extract data for specific populations such as pediatric patients, and additionally ADE data from international (EU, UK, and Japan) drug labels to build additional, complementary databases. To the best of our knowledge, this is the only current, up-to-date, multinational ADE database extracted from drug labels.

## Results

### Fine-Tuning a BERT-model to Accurately Evaluate ADE Terms

To utilize a language model for this task, we first need to identify a broad list of possible ADE terms from the drug label text. A model would then predict if each of these terms is a true ADE term. As such, we systematically compiled a set of possible ADE terms extracted from each individual drug label as described in the “Extracting Potential ADE Terms from Drug Labels” section, using a combination of exact string matching against the MedDRA vocabulary dictionary and terms predicted by DeepCADRME, a neural network model trained for ADE term extraction (20).

Once the potential ADE terms have been identified, we next train a language model to evaluate the likelihood of a given potential ADE term being a true ADE for a drug. We used the 200 SPL labels provided by Demner-Fushman et al. to train, validate, and test this model. For this task, we considered two BERT-based language models - ClinicalBERT, a model trained on electronic health record notes, and PubMedBERT, a model trained on PubMed research article abstracts (16,22).

Next, we optimized the performance of these language models for this task by fine-tuning the text input we provide for each candidate ADE term. (23) A detailed description of the parameters we evaluated is provided in the “Optimizing Language Model Input String” section. We concluded that an input string of 125 words, with 87.5% of the words coming after the candidate term was optimal, and identified that substituting the candidate term with a common token “EVENT” and prepending the term and section source to the input label additionally increased the performance of the models.

Finally, we ran comparisons between the best performing ClinicalBERT and PubMedBERT models to determine the model to utilize in constructing the OnSIDES database. We evaluated the performance of both models on the validation/test sets from the Demner-Fushman et al. annotated labels that were not used in the training of the models, and on the official TAC 2017 evaluation script that was used to benchmark models submitted to the ADR extraction task track of the workshop. Additionally, we further benchmark the performance of these models against the models proposed for the TAC 2017 workshop task. (18)

### OnSIDES : A Comprehensive, Computational Resource of ADEs

To construct the OnSIDES database, we first extracted a set of candidate ADE terms from all of the available drug labels in SPL format for the fine-tuned PubMedBERT model to evaluate. After we have predicted the ADEs for each drug label, to construct the database, we mapped the drugs and predicted ADEs to standardized vocabularies, RxNorm and MedDRA Preferred Terms (PT), respectively. As of the February 2024 release, the OnSIDES database includes 3,610,120 ADEs and 114,322 Boxed Warning ADEs from 47,211 drug labels, which includes 3,233 unique drug ingredient combinations (1,873 single-ingredient drugs, 1,360 multi-ingredient drugs) and 4,510 unique ADEs. To the best of our knowledge, OnSIDES is the only up-to-date and multi-national database of drug-label ADEs. It is also the only database that incorporates multi-ingredient drugs. The latest version of OnSIDES is publicly available online at https://github.com/tatonetti-lab/onsides. As new drugs are approved and drug labels are revised constantly, we have developed a computational pipeline to continually maintain and update the database quarterly and reflect the latest up-to-date knowledge available on ADEs, also available in the repository.

### OnSIDES-PED : Supplementary Resource of Pediatric ADEs

In the OnSIDES database, we compile ADEs listed on drug labels for the general population. However, there are specific populations that are known to be at higher risk for ADEs when taking certain medications. While the risk of serious ADEs is greater for pediatric patients, our knowledge is limited, with few RCTs being conducted on these populations given the restricted population and ethical concerns (24). Our lab’s previous efforts in constructing a database for pediatric ADEs, KidSIDES, focused on extracting ADE signals from spontaneous reporter databases whilst considering developmental stages (11). Intended as a database to be used alongside both the OnSIDES and KidSIDES databases, we constructed a database of drug label-extracted ADEs specific to pediatric patients.

To construct this database, we follow the same steps we describe for OnSIDES with slight modifications in the preprocessing steps. First, we identify the drug labels with any potential pediatric-specific ADE mentions in the “Specific Populations” section by extracting the subsections in these sections that mention the words “pediatric” / “child”. With this filtering, we extract 25,453 drug labels that include potential pediatric-specific ADE mentions in either of these two sections.

Here, as there exists no reference dataset of annotated drug labels, we manually annotate 200 randomly sampled drug labels to fine-tune and evaluate the fine-tuned PubMedBERT model. The steps taken to conduct the manual annotation is described in the “Manual Annotation of Drug Labels” section. Then, we follow the same input string construction methods and apply the BERT model fine-tuned for OnSIDES, but we optimize the model by re-computing the prediction cutoff for the pediatric ADE mentions. Here, we find that the adapted model achieves an overall accuracy of 0.64, precision of 0.61, and AUROC of 0.66 in extracting pediatric-specific ADE terms from the aforementioned sections, suggesting the flexibility of adapting this method to specific conditions. We use this modified method to construct a large pediatric-specific ADE database of 359,213 drug label-ADE pairs from 1,161 unique drug ingredient combinations, 1,526 unique ADEs, and 20,014 drug labels derived from the drug labels’ special populations section.

### OnSIDES-INTL : Supplementary Resource of ADEs Extracted from International Drug Labels

We have additionally generated OnSIDES-INTL, a complementary resource to OnSIDES, which has been extracted and compiled from European, British, and Japanese drug labels. The EU, UK, and Japan represent some of the largest pharmaceutical markets in the world outside of the US, with each having individual drug regulatory processes independent to that of the US (25–26).

To generate these databases, we adapt the OnSIDES method to best fit the respective drug label formats. We first download the raw drug label data, parse the labels’ text, and extract the ADE relevant sections. There are two main categories in which the ADEs are presented - free text and in tables. For the free text, we utilize the PubMedBERT model fine-tuned on the FDA drug labels. For the structured or tabulated ADE text, we extract exact matched strings from the tables. As the Japanese drug labels are not in English, we use the MedDRA-J (the Japanese MedDRA vocabulary) to extract terms from the tabulated data, and machine translate the free text using GPT-4 into English to run the OnSIDES model on.

Similarly to OnSIDES-PED, as there exists no reference dataset of annotated drug labels, we manually annotate 200 randomly sampled drug labels to fine-tune and evaluate the method. The procedure is described in the “Manual Annotation of Drug Labels” section. To fine-tune the model, we randomly selected 100 of these annotated drug labels to compute the optimal prediction cutoff for the model output for each section type, and used the remaining 100 drug labels to evaluate the model’s performance. For the tabulated data, we include all of the extracted exact matches, as the extracted terms are not assigned an ADE term likelihood probability.

After this evaluation, we run this process for all of the drug labels publicly available for each nation/region. By doing so, we construct additional databases with 108,958, 705,917, and 237,827 unique drug-ADE pairs from 1,048, 9,003, and 6,541 drug labels, with 929, 1,889, and 1,441 unique drug ingredient combinations and 3,570, 4,848, and 3,296 unique ADEs from European, British, and Japanese drug labels, respectively. Furthermore, by using international drug labels, we incorporate ADE data for an additional 1,777 unique drug ingredient combinations not found in OnSIDES. The database is presented in a format mirroring the OnSIDES database, and will be regularly updated simultaneously as part of the data repository.

**Figure 1:**
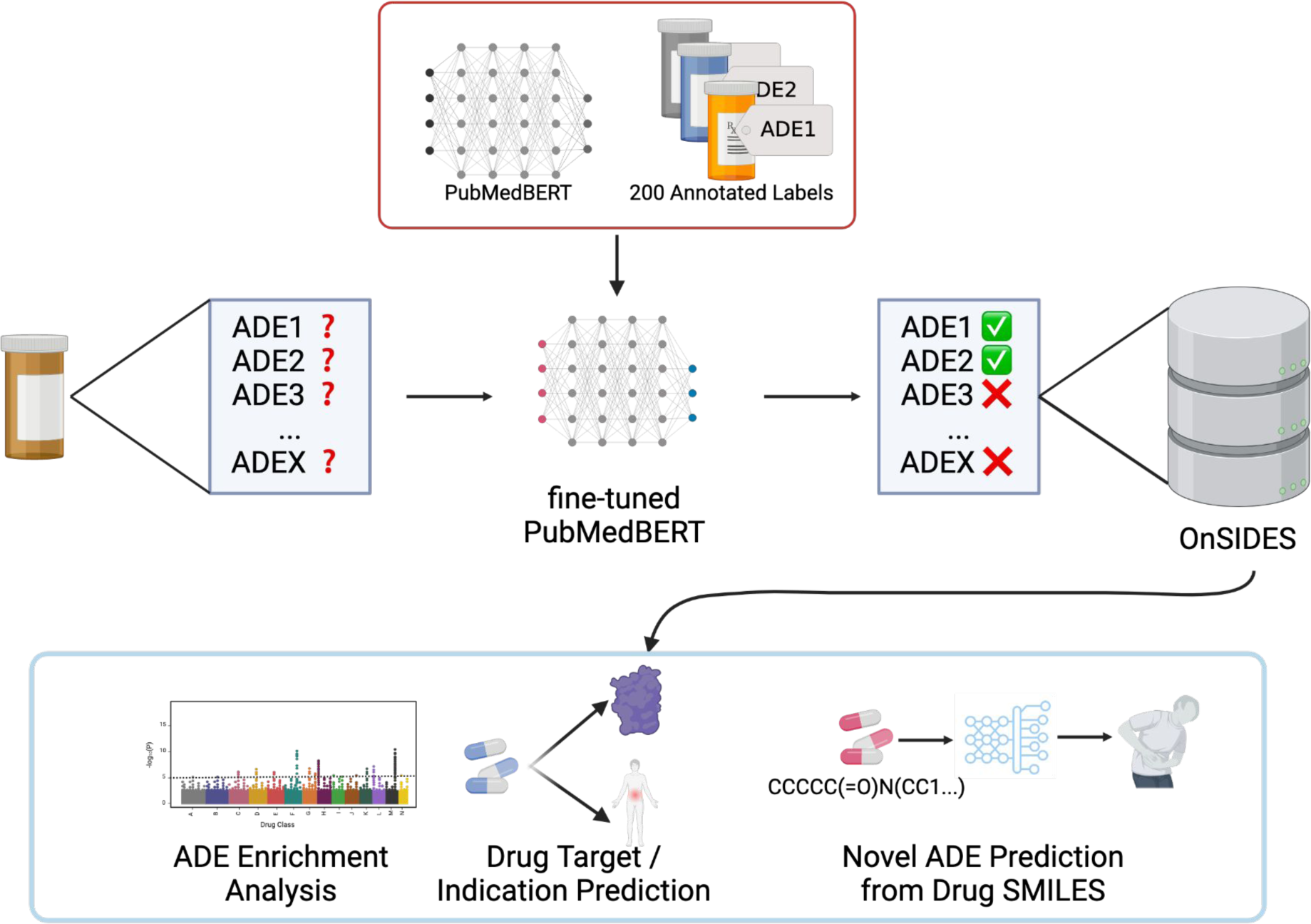
Graphical Representation of the OnSIDES Database Generation Process

### Interpreting the OnSIDES Prediction Model

Given the complex architecture of BERT models, it is often difficult to interpret the results they output. However, understanding the predictions that a given model makes is important in validating the model’s usability. We applied the SHapley Additive exPlanations (SHAP) method across each of the individual MedDRA System Organ Classes (SOCs), and computed local and global SHAP explanations for the ADEs within each class. Each explanation contains drug label text with overlaid importance weights as well as a dendrogram-fitted bar chart with the most important clusters of tokens, with red presenting positive influence and blue presenting negative influence.

We present examples of this method, the first of which is the explanation for a positively-labeled drug label for the ADE “thrombocytopenia”, in which the prepended token of the ADE name is the most significant predictor, lending support to the hypothesis that its inclusion improves predictive performance. (Figure 2A) The token “EVENT”, which is substituted for the ADE name throughout the text, also consistently acts as a strong positive predictor. Of note, this effect is influenced by the “EVENT” token being surrounded by numerical tokens, which is indicative of a tabular structure in the original drug label. Furthermore, an additional example shows an explanation for a negatively-labeled drug label for the ADE “tic” (Figure 2B). “Tic” refers to a neurological condition characterized by repetitive muscle movements, but its short name means that it can be erroneously identified in a drug label string. This is the case in this example, where the ADE name appears in the term “urTICaria”. SHAP negatively weights the prepended term and appears to propose an alternative hypothesis of what the drug label’s ADE can be by negatively weighting tokens related to “rash”. Examples of global explanations for MedDRA SOC categories have been added to (Supplemental Material 1).

**Figure 2:**
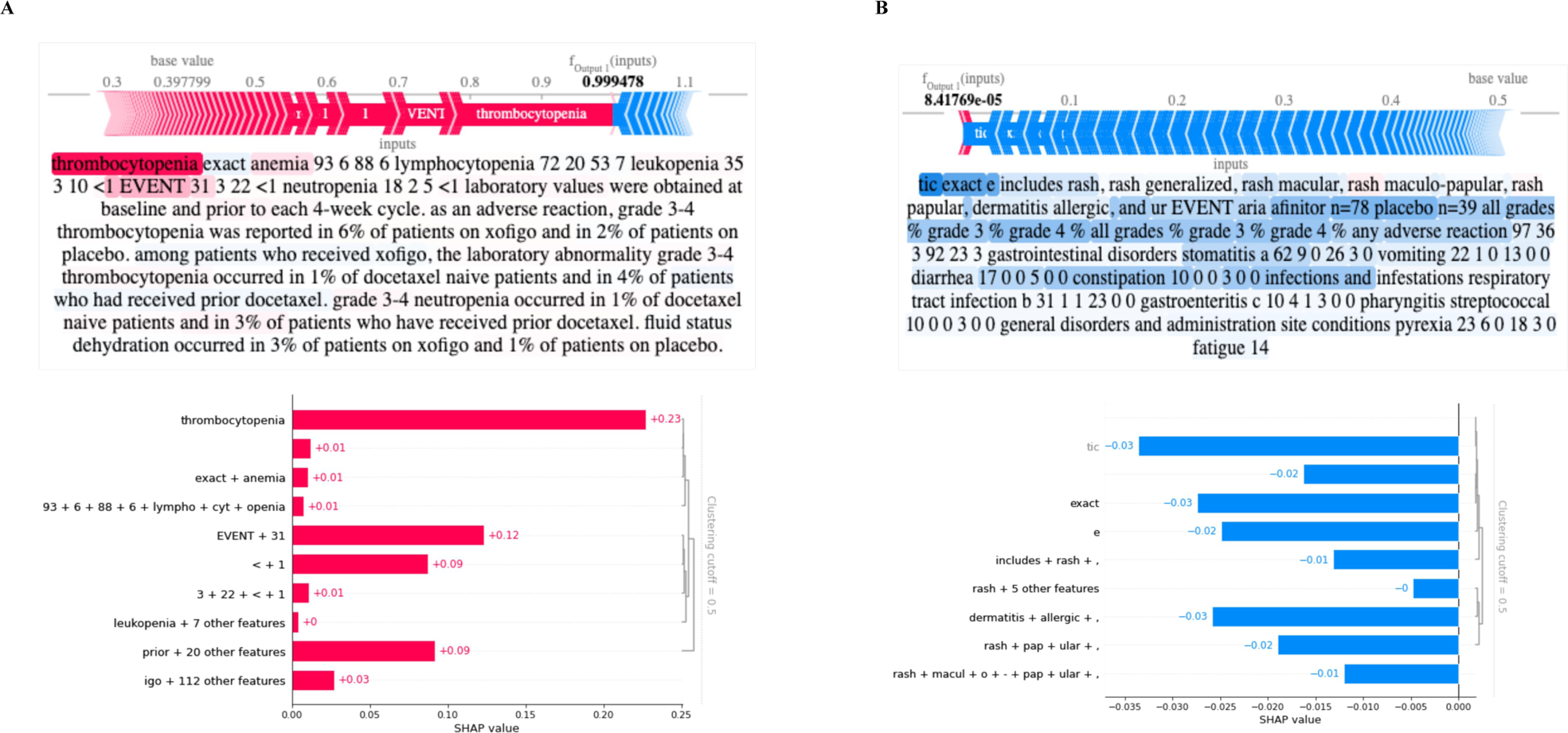
Explanation of OnSIDES Model Predictions using SHAP Values. A. SHAP explanations for a drug label positively indicated for the adverse effect thrombocytopenia. The top visualization shows the label text with contributions to the BERT model’s prediction indicated through SHAP values. Tokens shaded in red contribute to a higher prediction of adverse drug events, while those in blue suggest the opposite. Directly below this, a bar plot showcases the top features from that sample. Tokens are grouped based on their SHAP values with a clustering cutoff set at 0.5 to determine prominent clusters. The initial adverse event term thrombocytopenia and the substituted EVENT term are strongly positive features. The EVENT term appears between several numbers indicating that it is embedded with a tabular structure. B. Similar visualizations for a drug label negatively indicated for the adverse effect tic. The token tic inaccurately matches the middle of the word “uriTICaria”, which refers to a non-related condition. Many negatively contributing SHAP tokens seem to be related to rash, suggesting an alternative adverse effect of the drug than the one predicted.

### Multi-National Comparison of ADEs in Drug Labels

To stratify inter-national differences in ADEs listed on drug labels, we conducted a qualitative comparison of the OnSIDES and OnSIDES-INTL databases. To do so, we standardized the data across the databases by mapping the drug RxNorm identifiers to ATC 1st level (ATC1) terms and the ADE MedDRA PT identifiers to MedDRA SOCs, and conducted an analysis of the drugs and ADEs across each data source.

First, we compared the number of unique ADEs listed for drugs within each ATC 1st level drug class by each data source (Figure 3A). While we observed similar distributions in the number of ADEs across drug classes for all three data sources, we found that the EU had the fewest number of unique ADEs across all drug classes. Additionally, we found that drugs in the “antineoplastics and immunomodulators” class had the largest number of listed ADEs while the “antiparasitics and insecticides” class drugs had the fewest consistently across all data sources.

**Figure 3:**
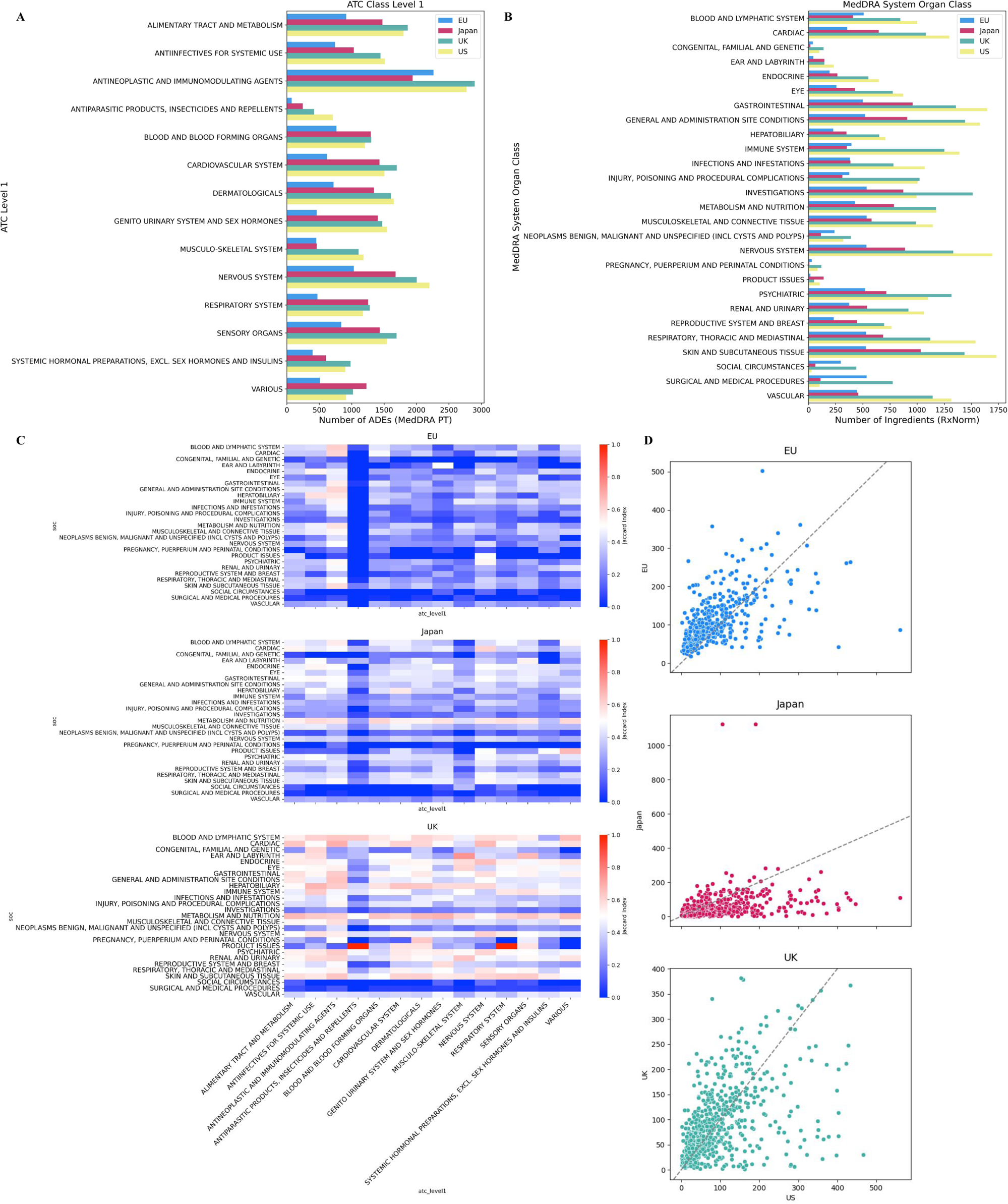
Overview of OnSIDES with a Comparison against OnSIDES-INTL (EU, Japan, UK) datasets A. Number of ADEs for drugs within each ATC Level 1 group, split by data source. Each bar represents the count of all unique ADEs (MedDRA PT) within the high-level group (ATC Level 1). B. Number of drugs having ADEs within each MedDRA SOC. Each bar represents the count of all unique drugs (ATC5) that have labeled ADEs (MedDRA PT) which belong to the ADE group (MedDRA SOC). C. Overlap in drug-ADE pairs between OnSIDES and OnSIDES-INTL. For each high-level group (ATC Level 1 and MedDRA SOC), we identified all unique drug-ADE pairs coming from each source, and the bin color represents the Jaccard index of the drug-ADE pairs between the two sources. D. Comparison of the per-drug number of ADEs (MedDRA PT) between OnSIDES and OnSIDES-INTL.

Second, we summarized the distribution of ADEs across the various SOC groups. For each SOC, we identified all ADEs within the group, then counted the number of unique drugs which list any of those ADEs (Figure 3B). Again, we observed a similar distribution of unique drugs across all SOCs. However, the EU dataset had the fewest number of drugs with an ADE listed within an SOC in most cases. Overall, the SOCs listed on the fewest number of drug labels were “product issues”, “pregnancy and related conditions”, and “congenital, familial, and genetic disorders” while “general disorders and administration site conditions”, “gastrointestinal”, “skin and subcutaneous tissue”, “nervous system disorders”, and “vascular disorders” were present in the largest number of drug labels.

Additionally, we evaluated the extent to which the three data sources contain the same drug-ADE pairs. For each data source, we grouped both the drugs and ADEs by high-level terms and calculated the Jaccard index (number of shared drug-ADE pairs divided by the total number of drug-ADE pairs) as a measure of the overlap between drug-ADE pairs between datasets (Figure 3C). Overall, we found that the majority of the drug-ADE pairs were not exactly shared between sources. However, there was some notable overlap in sensory organ drugs between the US and UK, and also antineoplastic and immunomodulating agent drugs among the US, UK, EU and Japan. Generally, the ADEs within the SOC for social circumstances and surgical/medical procedures had the least common drug-ADE pairs across the databases. This varying degree of overlap provides insight into the cross-comparability between data sources for different drug classes and ADEs.

Finally, we compared the number of ADEs (MedDRA PT) listed for each drug between different data sources to determine if the different data sources differ systematically. For each drug, we counted the number of unique listed ADEs within each data source (Figure 3D). These results show that US drug labels tend to have more listed ADEs than that of other nations, although considerable variation exists between drugs. Drugs tend to have the highest degree (number of ADEs) in the US data, followed by the UK data, while the Japanese data tends to assign the lowest degree. Despite apparent bias and variation between drugs, the number of listed ADEs per drug is clearly correlated between different data sources. Overall, while considerable variation exists, these results also show considerable similarity between different data sources.

### Enrichment of Pediatric-Specific ADEs in Drug Labels

In addition to understanding inter-national differences in drug label descriptions of ADEs, we conducted enrichment analysis of pediatric-specific ADEs extracted in OnSIDES-PED compared to the general OnSIDES database, and the KidSIDES dataset, which constructed a database of pediatric-specific ADEs mined from a spontaneous reporter, to identify if there were any distinct characteristics in each of these sources. To conduct a comparison between the KidSIDES, OnSIDES-PED, and OnSIDES databases, we standardized and grouped the drugs and ADEs across the databases by mapping the drug RxNorm identifiers to ATC1 terms and ADE MedDRA PT identifiers to MedDRA SOCs.

First, we conducted a comparison between the number of ADEs found for drugs in each ATC1 class for each dataset (Figure 4A). As expected by the spontaneous-reporter derived nature of KidSIDES, we found that there were significantly more ADEs in each ATC class of drugs when compared to OnSIDES/OnSIDES-PED. Next, we conducted a comparison between the number of drugs that have ADEs within each MedDRA SOC class. [Figure 4B] Again, KidSIDES had many more drugs in each category, highlighting the limited description of pediatric-specific ADEs in drug label text.

**Figure 4:**
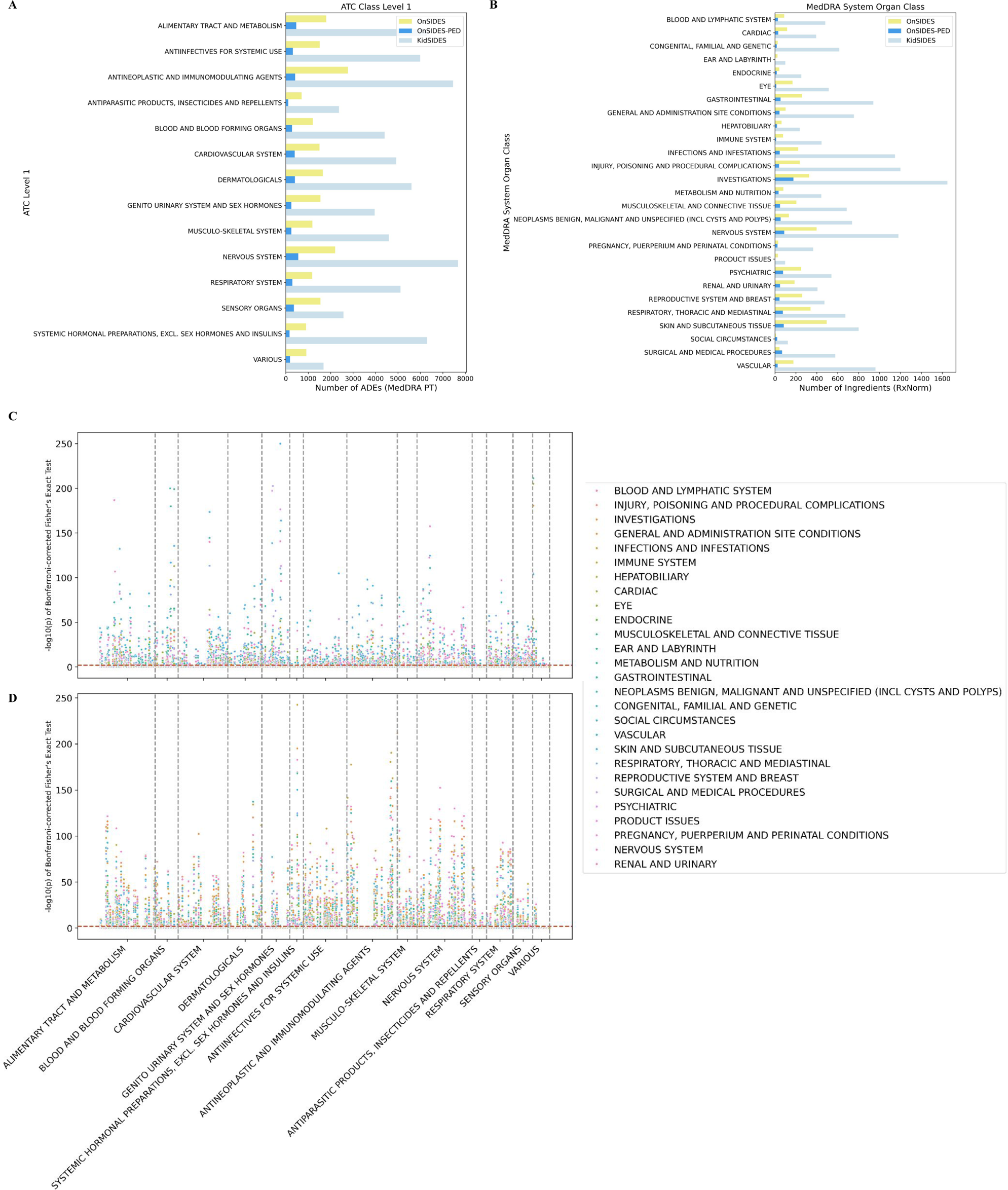
Comparison between OnSIDES, OnSIDES-PED, and KidSIDES A. Number of ADEs for drugs within each ATC Level 1 group, split by data source. Each bar represents the count of all unique ADEs (MedDRA PT) within the high-level group (ATC Level 1). B. Number of drugs having ADEs within each MedDRA SOC. Each bar represents the count of all unique drugs that have labeled ADEs (MedDRA PT) which belong to the ADE group (MedDRA SOC). C/D. A Manhattan Plot of the Enrichment of the Number of MedDRA PT Adverse Events for all Drug Classes (at the ATC Level 4). In C, we visualize the enrichment of OnSIDES-PED vs. OnSIDES. In D, we visualize the enrichment of OnSIDES-PED vs. KidSIDES. The markers are colored by MedDRA SOC.

Additionally, we conducted an enrichment analysis to identify any drug classes enriched with a certain adverse event type in the pediatric datasets by statistically testing for the number of mapped MedDRA PT terms for each drug class compared to all of the possible PT terms in each SOC. We compared OnSIDES and OnSIDES-PED [Figure 4C], and OnSIDES-PED and KidSIDES [Figure 4D]. In the comparison between OnSIDES and OnSIDES-PED, we observe an enrichment of a large number of “Skin and Subcutaneous Tissue” and “Gastrointestinal” ADEs, and very few “Social Circumstances” and “Pregnancy, Puerperium and Perinatal Conditions” ADEs enriched. Between OnSIDES-PED and KidSIDES, we observe an enrichment of ADEs in the “Nervous System” and “General and Administration Site Conditions” MedDRA SOC classes, and a relative lack of ADEs reported in the “Ear and Labyrinth” and “Social Circumstances” classes in KidSIDES.

This analysis recapitulates general trends in the enriched occurrence of specific classes of ADEs suggested by prior research efforts in this domain (11,27). Of note, we found specific occurrences of ADE descriptions in the pediatric drug labels related to the delayed elimination of amoxicillin due to the incompletely developed renal function in neonates and young infants, and the increased susceptibility to suicidal ideation and behaviour for adolescents when taking antidepressants. Additionally, this analysis allows for further insight into broadly identifying, classifying, and validating novel or poorly understood ADEs that pediatric patients are at higher risk for.

### Predicting Drug Targets and Indications from Pairwise Drug-ADE Similarity Scores

The work of Campillios et al. suggested the similarity of two drugs’ side effect profiles is positively correlated with the probability that these drugs share a common target. (28) Furthermore, Yang and Agarwal showed side-effect profile similarities can also predict shared indications. (29) By applying a simplified version of these methodologies, we computed pairwise Tanimoto Coefficient scores, a metric of the drugs’ side effect profile similarities for 1,485 OnSIDES single-ingredient drugs. The scores were then z-score normalized. Drug target and indication data for these drugs were mapped from DrugBank and MEDI respectively, and we computed how many, if at all, targets and indications every drug pair shared. Then, we used the normalized Tanimoto coefficient scores to predict the shared drug targets and indications using logistic regression models. We repeated these steps for the 1,430 drugs in the SIDER database to benchmark the predictive quality of the OnSIDES data.

We found that there is a logistic relationship between both the proportion of shared drug indications and shared targets and the shared side-effect profiles of drug pairs (Figure 5A). Additionally, we found that an OnSIDES-trained linear regression model performs moderately well in predicting whether a given drug has a specific drug target based on its side-effect profile, (AUROC = 0.667) and outperforms a model trained on SIDER (AUROC = 0.590). (Figure 5B) This suggests that the similarities between side-effect profiles found in drugs in the OnSIDES database may be used to predict novel drug targets of a given drug.

**Figure 5:**
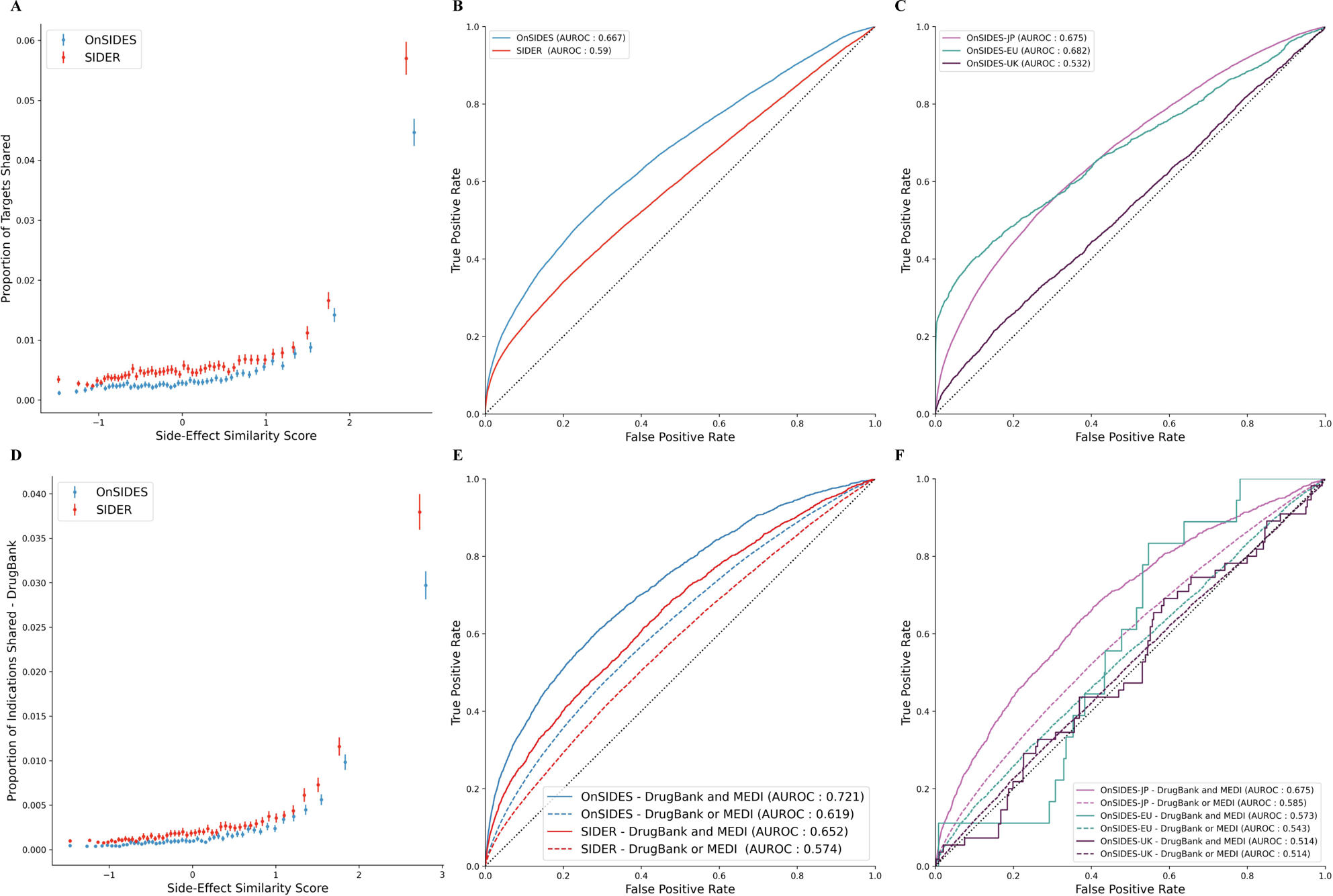
Predicting the shared drug targets and drug indications from drug pairs’ z-score corrected Tanimoto coefficient side effect similarity scores. A. The proportion of drug targets shared is correlated with the z-score corrected Tanimoto coefficient side effect similarity score. B. The OnSIDES-trained model outperformed the SIDER-trained model in predicting drug targets shared between pairs of drugs. C. Models trained on the OnSIDES-INTL (UK, EU, and Japan) databases also perform moderately well in predicting shared drug targets. D. The proportion of drug indications shared is correlated with the side effect similarity score. E. The OnSIDES-trained model outperforms the SIDER-trained model in predicting shared on-label (DrugBank) / off-label (MEDI) indications. Additionally, the predictions were more accurate when we took the indications listed in both DrugBank and MEDI. F. Models trained on the OnSIDES-INTL (UK, EU, and Japan) databases also perform moderately well in predicting shared on/off-label drug indications.

Next, we evaluated whether the OnSIDES and SIDER datasets could be used to predict the drug indications from DrugBank, MEDI, or the indications listed in both of the databases. We found that the OnSIDES-trained model was able to predict drug indications from each database, and also outperformed SIDER (Figure 5C). We further found that the OnSIDES dataset performed best in terms of predicting the existing therapeutic indications that were found in both DrugBank and MEDI (AUROC = 0.77) and better than SIDER in these predictions also (AUROC = 0.715), suggesting that these models perform well in predicting indications of high confidence (Figure 5D). This suggests that OnSIDES could be used as a potential resource for drug repurposing applications, identifying existing drugs that may be used for novel therapeutic indications, and to identify potential drug off-targets.

### Training ChemBERT Models on OnSIDES to Predict Population-Level ADEs

Significant advances in chemoinformatics and the application of Artificial Intelligence to drug discovery and drug repurposing tasks have increased the potential for successful in-silico driven development of new, effective medications for a variety of indications (30). The identification of potential toxicities that novel compounds may cause to patients is of particular importance, and may allow for safer and more effective clinical trials. As such, an in-silico model to predict ADEs directly from chemical compound structures, such as SMILES string texts, may contribute significantly to these efforts. Here, we trained and compared a number of chemoinformatics BERT models on OnSIDES to predict ADEs directly from chemical compound SMILES strings.

1,143 single-ingredient drugs from OnSIDES were converted to SMILES strings. We predicted whether these drugs are known to cause any of the 10 severe ADEs selected, which were mapped (Supplementary Table 3) to MedDRA HLTs (High-Level Terms). Using this data, we fine-tuned Chemprop, a graphical neural network, and 2 versions of ChemBERTa, a transformer model pre-trained on PubChem. (31–33) Additionally, we compared both random and scaffold based train/test split stratification.

Across these models, we achieve a test-set AUROC of 0.620-0.745 per ADE (Figure 6). We find that neither Chemprop nor ChemBERTa perform consistently better across all 10 ADEs. Scaffold-stratifying reduced performance, resulting in a best model test-set AUROC of 0.548-0.719 per ADR. In particular, ADEs 4 and 6 in particular drop significantly when scaffold stratification is applied, suggesting that their initial performance with random-split was confounded by train/test information leakage. Additionally, we hypothesize this dataset may not be large enough to achieve optimal performance in training deep learning models for this task. The incorporation of additional data, such as simulated data, may improve performance. We found that our OnSIDES-based HLT prediction models achieved performance on-par with previous work predicting less-granular SOC.

**Figure 6:**
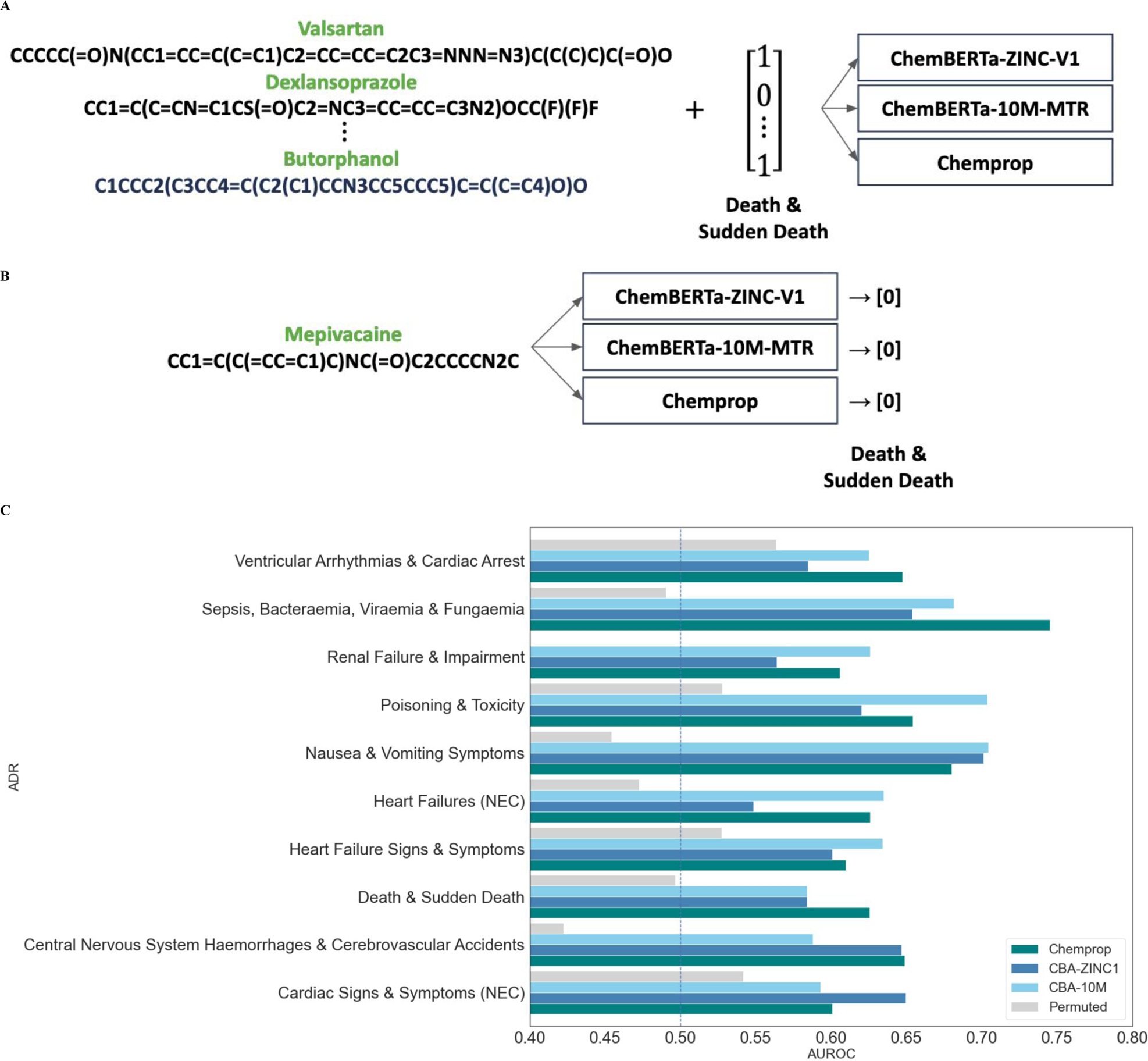
Overview of the SMILES-ADE Prediction Model Training and Testing. A. The models are trained using the SMILES strings of OnSIDES drugs and a binary vector indicating the presence of a particular serious ADE for that drug. B. The trained models are used to predict whether a testing set drug has the ADE listed on the drug label. C. The models achieve test-set AUROC of 0.620-0.745 per ADE, with no particular model demonstrably outperforming the others.

## Materials and Methods

### Extracting Potential ADE Terms from Drug Labels

In order to train and develop the model to construct the OnSIDES database, we downloaded a total of 46,686 FDA SPL drug labels from the NIH NLM DailyMed website (https://dailymed.nlm.nih.gov/dailymed/spl-resources-all-drug-labels.cfm). The SPLs are publicly available as individual XML files which are collated as a zipped file, and the resource is updated periodically (daily, weekly, or monthly). We further used a set of 200 SPL labels manually annotated for ADEs to train and validate the model (19). These labels are publicly available at (https://bionlp.nlm.nih.gov/tac2017adversereactions/), and are provided as XML files, along with an additional 2208 unannotated drug labels.

The SPL format of drug labels is a document markup format approved by the XML-based HL7 clinical and administrative data standard utilized by the FDA to present relevant pharmaceutical information text in a standardized manner (34). All entities such as pharmaceutical companies that submit drug approval applications to the FDA are required to submit documentation in this format, thereby allowing the FDA to receive and provide information to healthcare professionals and patients in a consistent manner (35). The SPL format incorporates a broad range of sections including the drug’s indications, clinical pharmacology, contraindications, warnings, and ADEs (36). In the FDA SPL drug labels, the information related to ADEs is primarily located in the “Adverse Reactions” (AR), “Boxed Warnings” (BW), and “Warnings and Precautions” (WP) sections of the labels. We primarily utilized the AR and WP sections, and further extracted serious warnings from the BW section as a separate model / table.

We evaluated the candidate ADE term identification performance between two different term extraction methods - exact string matching and combining exact string matches and extraction using DeepCADRME, a neural network model trained for ADE term extraction (20). Exact string matching was conducted using the MedDRA vocabulary dictionary as reference, and extracting any strings that were identical to any MedDRA PT and LLT terms. DeepCADRME is trained on the Demner-Fushman et al. annotated label dataset that we also utilize in the model training, and uses text sequences to build N-level contextualized embeddings to annotate ADE terms. The exact string matching-only extraction achieved a F1 score of 0.890 and an AUROC of 0.880, while the combined method of extraction achieved a F1 score of 0.891 and an AUROC of 0.900 when tested on a test dataset of 100 labels using a PubMedBERT model. Further statistics are available in (Supplementary Figure 1). As such, while there was only a marginal improvement in performance, we conducted extraction using both to construct the OnSIDES database. In addition, this suggests that if a user does not have results extracted from DeepCADRME available, the performance of the model is not hindered significantly.

### Optimizing Language Model Input String

In order to improve the performance of the language models we have selected for this task, we evaluated different methods of input string construction to determine the optimal input string to extract from a drug label section to predict the likelihood of a candidate ADE term we identified in the previous step being a true ADE for a given drug.

First, hypothesizing that the model could learn ADE context generally, we examined whether replacing the potential ADE term with a common term “EVENT” would improve the performance of the language models. We compared this to a nonsense, unmapping term as a control. Additionally, we tested whether prepending the potential ADE term or the name of the drug label’s section it was extracted from to the example string would improve the performance of the models. We iterated over the combinations of these parameters, and found that substituting the term with the common token and prepending the term and section source to the input label resulted in the best performance for both the ClinicalBERT and PubMedBERT models. With this combination of parameters, we achieved F1 Scores of 0.861 and 0.874, and AUROC Scores of 0.896 and 0.899, respectively for the ClinicalBERT and PubMedBERT models. The detailed performance metrics for the various combinations of parameters is described further in (Supplementary Figures 2). We found that prepending the source term most significantly improved the performance of the model compared to the baseline input, but the combination of all parameters described above performed the best.

Furthermore, we hypothesized that more context may lead to better performance and that context that comes before the ADE mention may have different value than context that comes after. We conducted experiments to examine whether the performance of the models would vary depending on the length of the input string that was extracted from the drug labels, or the proportion of the extracted string before and after the ADE term in question. We adjusted the length of the input string between 30 and 250 words, which was our maximum in using the language models. We found that the 125 word input string performed best, closely followed by the 60 word input string (Supplementary Figure 3A). This suggests that a considerable amount of context aids performance, but too long of an input string may dilute the important context with unrelated noise. Additionally, the proportion of string before and after the potential ADE term with which the model achieved peak performance was when 87.5% of words came after the term, followed by 75% after the term (Supplementary Figure 3B).

### Comparing PubMedBERT and ClinicalBERT

To determine the final model to be used to construct the OnSIDES resource, we compared the best performing models between PubMedBERT and ClinicalBERT. Both share similar architectures, broadly leveraging a large corpus dataset of relevant text to build a pre-trained model, which can be adapted more specifically to undertake downstream biomedical and clinical NLP tasks. These models were chosen as they are trained on biomedical language similar to that found on drug labels, and were demonstrated to perform well on a variety of relevant NLP tasks (37). We evaluated the performance of both models on the Demner-Fushman et al. annotated label set, with both a standard train/validation/test 80/10/10 split used to train the model, and the TAC 2017 evaluation script. The PubMedBERT-based model achieved a modest performance improvement over the ClinicalBERT-based model, and notably it performed more consistently across the two different evaluations. The detailed performance metrics for each model and evaluation set are detailed in (Supplementary Figure 4A). Furthermore, we benchmark the performance of the PubMedBERT model to all of the computational models proposed for the TAC 2017 workshop, and the ADE terms listed in the SIDER database. We find that our model outperforms all TAC 2017 models and the SIDER database as presented in (Supplementary Figure 4B).

### Applying the Model to International Drug Labels

In addition to constructing an OnSIDES database from FDA drug labels, we leverage the method to expand the database to incorporate ADE data found in drug labels of other nations/regions. As such, we adapted our method to extract ADEs from drug labels from the EU, UK, and Japan to generate additional computable resources to comprehensively study ADEs. In each country/region, the EMA (European Medicines Agency), MHRA (Medicines and Healthcare products Regulatory Agency), and PMDA (Pharmaceuticals and Medical Devices Agency) are the respective regulatory bodies overseeing pharmaceutical product approval (38–40).

We obtained the corresponding drug label data from the following: (i) the EU’s 1,736 SmPC (summary of product characteristics) files (38); (ii) 9,334 drug SmPC files from the UK’s EMC (electronic medicines compendium) (41); and (iii) 24,802 Japanese drug labels from KEGG MEDICUS (42). All of these files are in a standardized format by country/region, and contain similar information as in the FDA SPL drug labels, including drug composition, contraindications, indications, ADEs, and warnings for specific populations. They are all publicly available, regularly updated sources, and the EU files are in PDF format while the UK and Japan drug data were constructed from their individual drug webpages as HTML files. We have processed these drug label files into individual standardized, structured files for public use in further model training / methods development. Data availability is described below.

To develop this international database, we utilized a combination of rule-based extraction and PubMedBERT-based NLP extraction methods. While the FDA SPL drug labels primarily list the ADEs in a free text format, drug labels from other regulatory agencies format the ADE section in structured lists, tables, or a combination of the three formats. As such, we adapted our extraction method to optimize for and accurately reflect the label sections we were extracting from (43). The rule based extraction, which was used for extracting ADEs written in a structured list or tabular format, consisted of extracting System Organ Class (SOC) - ADE - ADE Frequency triplets systematically from the lists or tables using exact term matching using MedDRA LLT (Lowest Level Terms)/PT and SNOMED CT condition terms, which were standardized to MedDRA PT codes. For the free text sections, we used the method we developed to evaluate the ADEs in FDA drug labels. While the EU and UK drug labels are in English, the Japanese drug labels are not. To overcome this issue, we utilized rule based extraction using MedDRA-J, a standardized translated MedDRA vocabulary, and machine translated the free text sections to apply the OnSIDES method to. Finally, in order to validate the performance of the developed extraction method, we manually reviewed and annotated 200 randomly sampled drug labels each from the EU, UK, and Japan.

### Manual Annotation of Drug Labels

To conduct validation on the non-FDA drug labels and the pediatric sections of the FDA drug labels, we conducted manual annotation of general ADE mentions in the UK, EU, Japan drug labels and the pediatric sections in the US drug labels following a similar approach to (19). First, we randomly sampled 200 labels for each drug label type. Next, we extract the relevant section of the drug label, and verify that the section we have extracted contains relevant ADE mentions. While general ADE descriptions were found to be standardized across drug labels / label types, pediatric ADE descriptions were often found nested in the general ADE sections, or did not exist anywhere in the label text due to the lack of information. Additionally, some pediatric ADE descriptions consisted only of a statement similar to “this medication has not been assessed for safety in pediatric patients”. To construct a relevant, evaluable dataset, we drop all labels with no relevant information available, and resample to create datasets of 200 labels with relevant information. We then manually review and annotate each drug label section text for all ADE mentions using doccano, a open-source text annotation tool (44). After annotation, we map all ADE mentions to standardized MedDRA terms. We use this manually annotated dataset to evaluate the accuracy of our model in the generated datasets. This data is also available in this project’s repository, further described in the “Data Availability” section.

### Explainability of fine-tuned PubMedBERT model

In order to better understand how the PubMedBERT model we have trained is determining whether a given term is an ADE, we utilize the SHapley Additive exPlanations (SHAP) method (45). The SHAP method is a game theory-based approach that explains model predictions by assigning individual features an importance value for a specific prediction, allowing for a better understanding of how individual features impact model decisions. The importance values for predictions can be computed in both local, single-sample contexts and global, whole set contexts. Here, we applied the SHAP method on OnSIDES predictions of ADEs from the drug labels in the test set to understand which string tokens contribute to positive and negative predictions. We grouped the ADEs by the 24 MedDRA System Organ Classes (SOC), and then computed local and global SHAP explanations for the ADEs within the class.

### Overview and Multi-National Comparison of ADEs using OnSIDES and OnSIDES-INTL

Previous studies have suggested that there is a significant inter-national difference in ADE listed on drug labels depending on the ADE in question and therapeutic area given the differences in drug safety regulation and drug approvals (46). As such, we sought to provide a qualitative overview of the OnSIDES database through a comparative analysis against the OnSIDES-INTL database. We first mapped the OnSIDES data from lower-level terms to higher-level terms to evaluate patterns in the different data sources. Low-level terms were coded as MedDRA PT for ADEs and RxNorm ingredients for drugs. Then, we mapped the drug ingredient RxNorm identifiers to Anatomical Therapeutic Chemical (ATC) 5th level (ATC5) and then mapped ADEs to the high-level MedDRA System Organ Class (SOC) and drugs to drug classes, coded as ATC 1st level (ATC1).

Firstly, we compared the number of unique ADEs listed for the drugs within each ATC1 drug class for each individual database. Next, we observed the distribution of ADEs across each MedDRA SOC group for each database. Then, we computed the Jaccard index for the drug-ADE pairs grouped by the high-level terms for each database to compare the overlap of drug-ADEs between each individual database. Finally, we compared the number of most granular (MedDRA PT) ADE terms listed for each drug ingredient terms between each database to determine if the databases differ systematically. Through this analysis, we identified drug and ADE classes with significant variation and similarity across databases.

### Applying OnSIDES : Prediction of Protein Targets and Drug Indications using Drug-ADEs similarities

As an example of the computational applicability of the OnSIDES database, we used the OnSIDES-derived pairwise side-effect similarity score dataset to build logistic-regression models using these scores as a predictor for the proportion and absolute number of shared targets and shared drug indications. Then, to benchmark the predictions, we compared it to the predictive performances of models built from the SIDER database-derived predictions.

To conduct this analysis, we mapped the drugs and ADEs between the OnSIDES and SIDER datasets using standardized vocabulary using OHDSI Athena mappings (47), and extracted datasets of drugs and conditions that were in both the OnSIDES and SIDER datasets. OnSIDES contains 1,485 unique single-ingredient drugs and 4,345 unique side effects, and SIDER contains 1,430 unique drugs and 4,251 unique side effects. Of these drugs and side effects, there are 1,026 common drugs and 3,181 side effects as mapped to standardized vocabularies. Then, we obtained drug target information from DrugBank, and drug indication information from DrugBank and MEDI, and mapped the OnSIDES and SIDER ADE datasets to the respective datasets (48–49). DrugBank primarily contains drug indications extracted from drug labels, while MEDI utilizes public internet sources to drug indications, and is a well utilized resource of drug indications. There are 1,474 and 1,347 drugs with indications listed in DrugBank, and 1,257 and 1,155 drugs with indications listed in MEDI for OnSIDES and SIDER respectively. Further mapping statistics provided in (Supplementary Material 3).

Finally, we used the constructed dataset to build logistic regression models, using the z-score normalized Tanimoto coefficient metrics for the drug pairs to predict the shared drug targets and indications of a given pair of drugs. The Tanimoto Coefficient is a metric to measure the similarity of two sets of features by computing the ratio of the number of common elements to the total number of elements (50). After computing these metrics, we applied a z-score normalization across each dataset by normalizing on the mean of each drug’s similarity scores to remove the bias of drugs having a higher similarity score on average.

### Applying OnSIDES : Comparison Chemoinformatics Methods for the Prediction of Population-Level ADEs from SMILES Strings

Additionally, to explore other potential applications of the OnSIDES database, we fine-tuned and compared chemoinformatics BERT models on OnSIDES to predict ADEs directly from chemical compound SMILES strings. This is an area of active interest, with different methods being proposed in recent years, given its potential utility in identifying chemical compounds with/without severe toxicity to humans during preclinical/clinical development. (33, 51)

To develop the SMILES-ADE prediction models, we first constructed the training/testing datasets from the OnSIDES database. 1,143 single-ingredient drugs from OnSIDES were converted to SMILES strings using PubChemAPI. We converted 10 severe ADEs (Supplementary Table 3) to MedDRA HLTs (High-Level Terms), which are more granular than the MedDRA SOC (System-Organ Class) terms predicted in the most relevant previous research study in this field. Additionally, we identified three different models of relevance, Chemprop, ChemBERTa, and ChemBERTa-10M-MTR, and fine-tuned each of these models to compare their performance. ChemBERTa is a transformer-based model of RoBERTa architecture (52) that takes SMILES strings as input and can be fine-tuned to predict various chemical and physical targets, while Chemprop is a graphical neural network. Ahmad et al. (32) found that Chemprop performed better than ChemBERTa for ClinTox classification, a drug-toxicity classification task. However, ChemBERTa was found to perform better than Chemprop on a number of other MoleculeNet benchmark tasks (51), suggesting that either model could be utilized. Additionally, we compared these models with random and scaffold based train/test split stratification.

## Discussion

### OnSIDES : Enabling large-scale, broad studies of ADEs

ADEs are a significant cause of morbidity, mortality and unnecessary additional medical treatment worldwide. Drug label information provided by regulatory agencies continues to be the gold standard of adverse drug event information available for individual drugs. Furthermore, the advancement of pharmacology-related computational methods has the potential to revolutionize drug discovery, clinical treatment and precision medicine, and enable accurate prediction of novel drug interactions, side effects, and therapeutic potentials. To realize this, the field requires high quality, standardized, and comprehensive datasets to train computational models on. However, no actively maintained comprehensive ADE databases exist, making it difficult to leverage computational tools to study ADEs broadly. As such, we present OnSIDES, a large-scale, machine-readable ADE database built by fine-tuning a NLP model, PubMedBERT, to accurately extract ADE terms from drug labels. Through the drug repurposing and chemoinformatics example projects presented here, we additionally show that OnSIDES can be used in a wide variety of pharmacovigilance and related applications. We believe that OnSIDES is a valuable resource that can be utilized to comprehensively study ADEs, and be further applied to different avenues of research to aid the advancement of precision pharmacology.

In generating OnSIDES, we present a method to fine-tune a large NLP model that has been pre-trained on general biomedical text for the specific task of classifying ADE-related terms given relevant context. To achieve optimal performance, we compared the performance of two pretrained NLP models, PubMedBERT trained on biomedical abstracts and article text and ClinicalBERT trained on a large corpus of electronic health record text. We fine-tuned both models by comparing different extraction methods, standard vocabulary-based exact string matching and neural network-based extraction, replacing the potential ADE term with different tokens, testing different input string lengths and different proportions of string before and after the ADE term, and adjusting the learning rates. We found that there was marginal improvement when we introduced neural network based extraction and that substituting the potential ADE term with the common token “EVENT” and prepending the term and section source to the input label improved both models’ performance further. Finally, we utilized a relatively long input string of 125 words split with 87.5% of the input string’s words coming after the term, suggesting that in this context a large amount of post-term context aids performance of the NLP model. While both models performed similarly well, we decided to use the fine-tuned PubMedBERT model as it achieved greater consistency in the multiple evaluations we conducted.

The final model trained for the OnSIDES database may be able to achieve strong performance of ADE term extraction from other types of text, and this approach of fine-tuning a large pre-trained language model for specific tasks can be adapted to be used for the extraction and classification of a specific class of biomedical terms from any relevant text.

### Limitations of OnSIDES

The OnSIDES database is constructed using computational extraction methods, with some side effects not extracted accurately, and some predicted adverse events being incorrect. We expect further methodological advances, both in potential ADE term identification and ADE term prediction, will improve ADE term extraction accuracy. Additionally, as we have exclusively extracted information from drug labels, there may be known ADEs that are not yet reflected on drug labels that may appear in other sources, such as adverse event reporting systems. Furthermore, as we use the MedDRA vocabulary as our starting point, we are unable to identify ADEs that are described in non-standard terms, which is a key component that hinders extraction accuracy. While we have strived to optimize the accuracy of drug-ADE pairs in the database, when conducting studies on specific drugs / adverse events, the most up-to-date drug labels should be referenced to verify information in OnSIDES.

Our additional databases, OnSIDES-PED and OnSIDES-INTL also face their own limitations in addition to those described above. OnSIDES-PED is currently compiled from ADE mentions in the BW, SP, and WP sections. As such, this does not include any general ADEs that pediatric patients may be affected by, that may be listed in the AR section alongside the adult ADEs. This is predominantly due to the fact that the pediatric mentions in this section are difficult to stratify systematically, and cause inaccuracies in the data. Additionally, it includes any information extracted from text describing pre-clinical trials, such as ADEs identified through *in-vivo* experiments conducted in juvenile rats, that may not directly occur in a human pediatric patient. Furthermore, the limited accuracy of the model may be improved by further adapted curation of the input format, rule-based filtering of unrelated matched terms, and the training of a model on the pediatric-specific mentions. For the OnSIDES-INTL database, we are limited to the currently active drug labels, as any previous versions of drug labels are unavailable in the public domain to the best of our knowledge. As we periodically update the database, we will be able to accumulate more data across all data sources and provide a more comprehensive picture incorporating more drug labels. Another limitation factor for this method is the machine translation accuracy of the OnSIDES-JPN labels. We hope to accumulate further annotated data to train/fine-tune specific models to optimize performance for each OnSIDES-INTL data table, and for non-English language texts.

Furthermore, we additionally maintain the OFFSIDES and TWOSIDES databases, which extract ADEs and drug-drug interactions respectively from the spontaneous ADE reporting system FAERS (FDA Adverse Event Reporting System). These are additional complementary resources that are available to be used alongside OnSIDES to comprehensively study adverse drug events at a large scale (Tatonetti, 2012).

### Future Directions

Here, we show that we are able to extract pharmacovigilance information with good accuracy from drug label data structured in a variety of different text formats using Natural Language Processing models. However, there is more pharmacologically relevant information nested in drug labels such as concomitant drugs, clinical trial results relevant to ADEs, pharmacogenomics, pharmacodynamics and pharmacokinetics information, and different ADEs noted for special populations of particular interest, that would be of great utility if standardized. Additionally, given the rapid development of NLP methods with the advent of Large Language Models, we expect extraction of information from free text to improve further, for example, ADEs for specific patient populations that are nested within general ADE-relevant text. We hope to further extend this work to extract and compile different data points by further adapting and developing this approach.

## Acknowledgements

This work was primarily supported by the National Institutes of Health (NIH), National Institute of General Medical Sciences (NIGMS) grant R35GM131905. Additionally, U.G, M.Z, K.L.B are supported by a NIH National Library of Medicine (NLM) grant T15LM007079, and H.Y.C is supported by the NIH NIGMS grant T32GM145440. The content is solely the responsibility of the authors and does not necessarily represent the official views of the NIH.

The graphical abstract (Figure 1) was created using Biorender.

## Author Contributions

Conceptualization: N.P.T

Methodology: Y.T, H.Y.C, N.P.T

## Database Development and Implementation : All Authors

Investigation and Visualization : Y.T., U.G, J.K., J.P., A.S., M.Z

Writing - Original Draft : Y.T, P.B., and N.P.T

Supervision : N.P.T

Funding Acquisition : N.P.T

## Data and Code Availability

All of the data, code, and models trained and generated to construct the OnSIDES database and all other complementing databases are available and maintained at https://github.com/tatonetti-lab/onsides. Any requests for additional materials can be made via email to the corresponding author.

