## Supplementary Materials for "OnSIDES (ON-label SIDE effectS resource) Database : Extracting Adverse Drug Events from Drug Labels using Natural Language Processing Models"

**Table 1 : Acronyms Used in this Publication**

ADE : Adverse Drug Events
AR : Adverse Reactions
ATC : Anatomical Therapeutic Chemical
AUROC : Area Under Receiver-Operating Characteristic curve
BERT : Bidirectional Encoder Representations from Transformers
BW : Boxed Warnings
EMA : European Medicines Agency
EMC : Electronic Medicines Compendium
FAERS : FDA Adverse Event Reporting System
FDA : Food and Drug Administration
MedDRA : Medical Dictionary for Regulatory Activities
MedDRA HLT : MedDRA High-Level Terms
MedDRA LLT : MedDRA Lowest Level Terms
MedDRA PT : MedDRA Preferred Terms
MedDRA SOC : MedDRA System Organ Classes
MHRA : Medicines and Healthcare products Regulatory Agency
NLP : Natural Language Processing
NIH NLM : National Institutes of Health, National Library of Medicine
PMDA : Pharmaceuticals and Medical Devices Agency
SHAP : SHapley Additive exPlanations
SmPC : Summary of Product Characteristics
SMILES : Simplified Molecular-Input Line-Entry System
SNOMED CT : Systemized NOMenclature of MEDicine, Clinical Terms
SP : Special Populations
SPL : Structured Product Labels
WP : Warnings and Precautions
XML : Extensible Markup Language

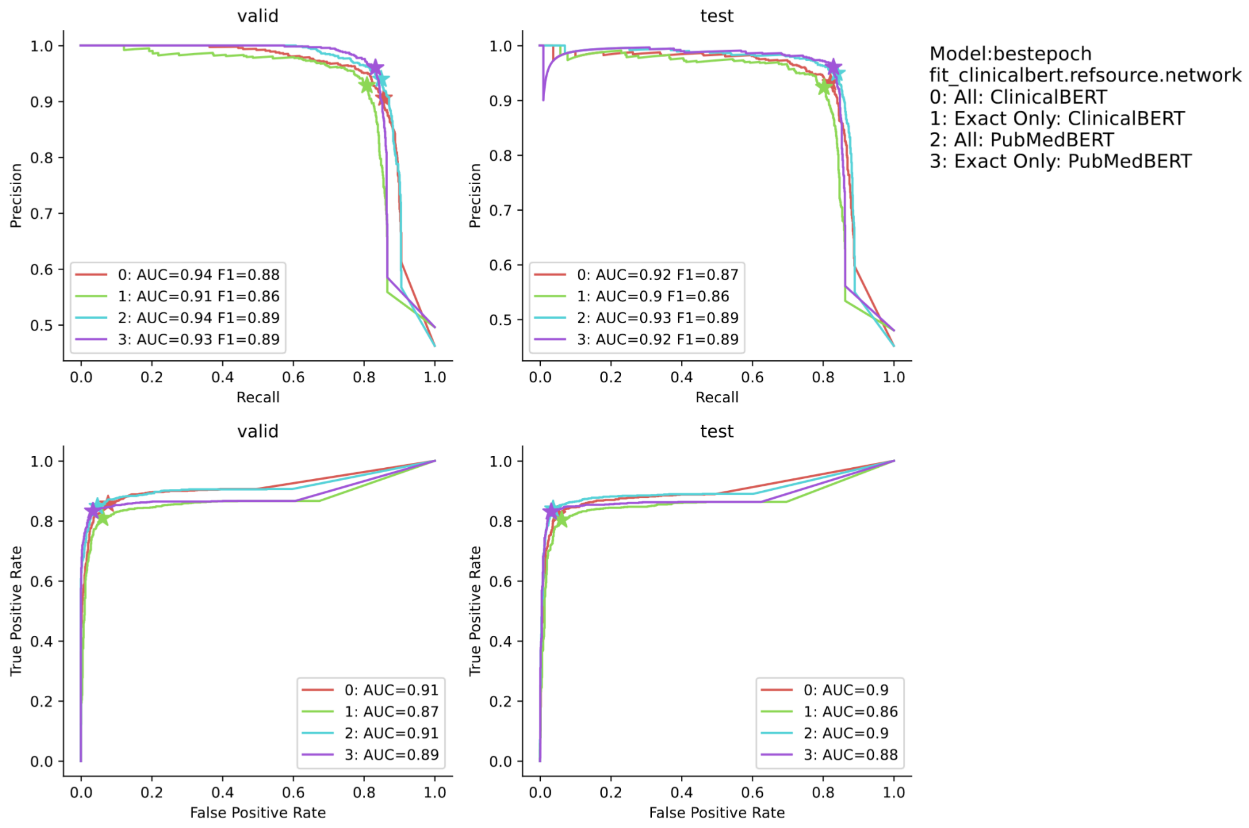


**Supplementary Figure 1: A Comparison of Input String Types for ClinicalBERT/PubMedBERT models.**

A comparison of the ClinicalBERT/PubMedBERT models' performance when trained on both DeepCADRME-extracted and exact-matched strings, and only exact-matched strings. We see a marginal improvement in performance when the either model is trained on both the DeepCADRME-extracted and exact-matched strings.

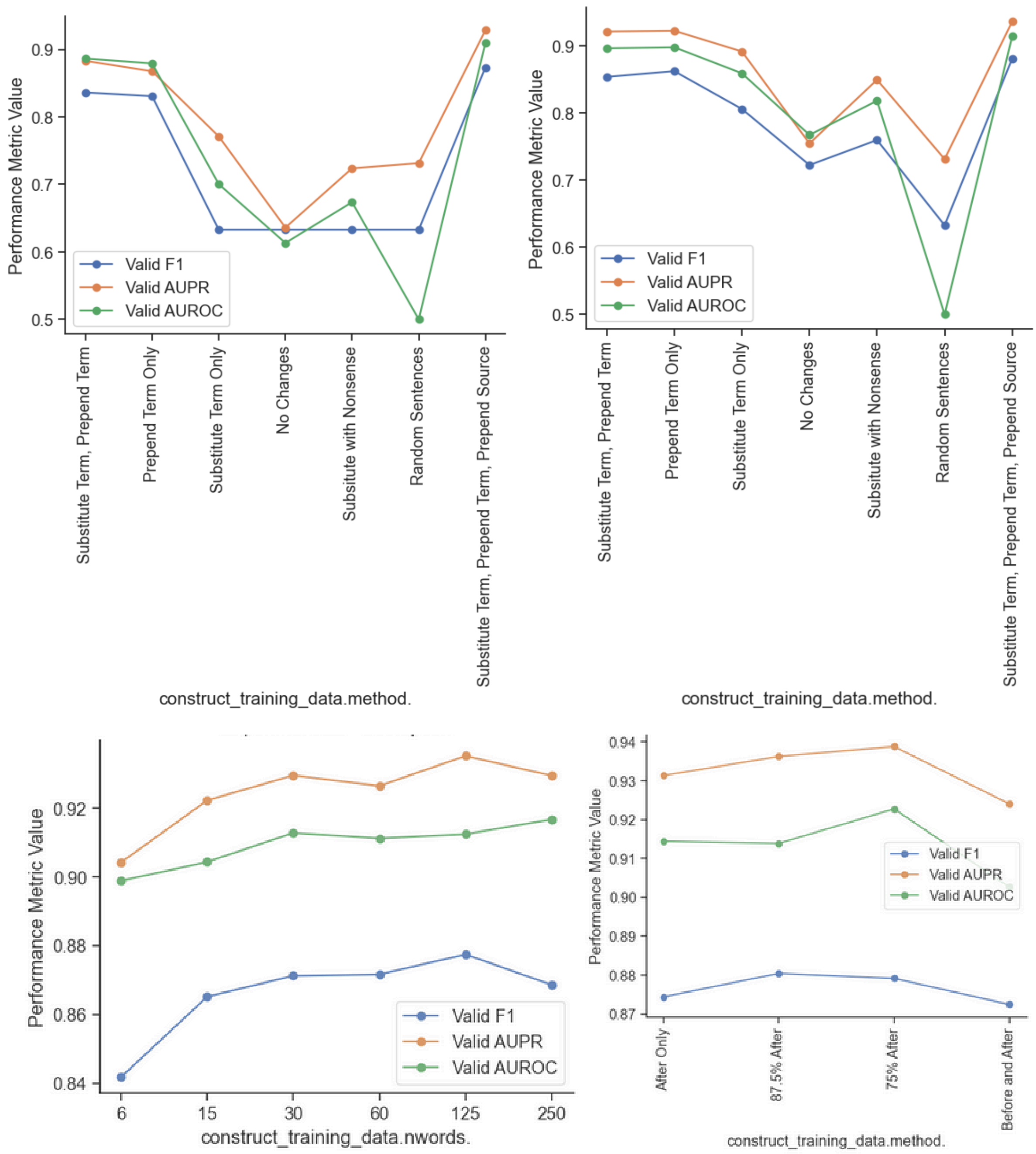


**Supplementary Figure 2 : A Comparison of Input String Format for ClinicalBERT / PubMedBERT models.**

A comparison of the ClinicalBERT (left)/PubMedBERT (right) models' performance when trained on both DeepCADRME-extracted and exact-matched strings, and only exact-matched strings. We see a marginal improvement in performance when the either model is trained on both the DeepCADRME-extracted and exact-matched strings.

C. The model performs best when we use a 125 word input string (Test AUROC : 0.900 / F1 : 0.877), closely followed by the 60 word input string (Test AUROC : 0.900 / F1 : 0.867).

D. The model achieved peak performance when 87.5% of words in the string came after the term, followed by 75% after the term in question.

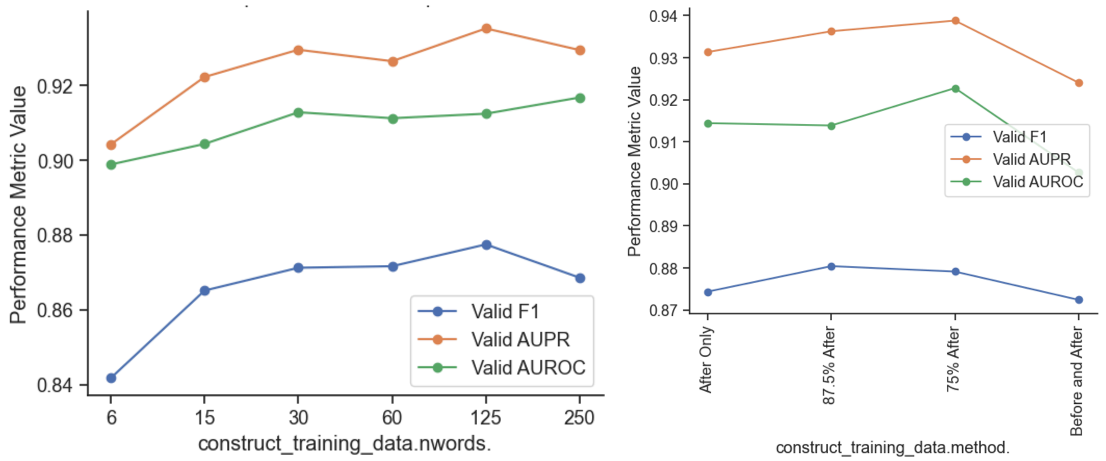


**Supplementary Figure 3 : Comparison of Prediction Performance with Differing Input String Lengths and Pre/Post String Splits.**

Figure A. The model performs best when we use a 125 word input string (Test AUROC : 0.900 / F1 : 0.877), closely followed by the 60 word input string (Test AUROC : 0.900 / F1 : 0.867).

Figure B. The model achieved peak performance when 87.5% of words in the string came after the term, followed by 75% after the term in question.

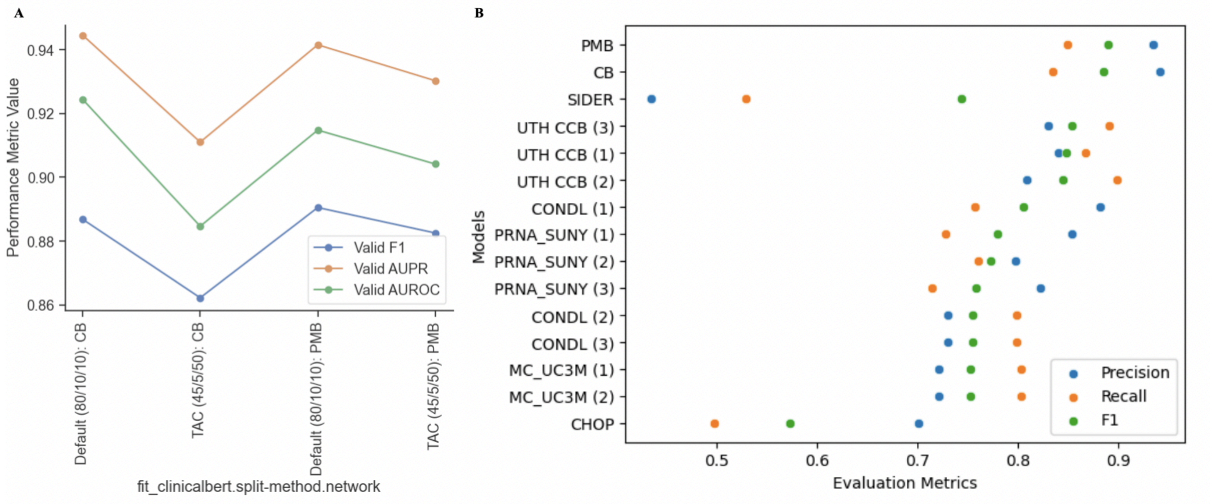


**Supplementary Figure 4 : Comparison of Prediction Performance between ClinicalBERT, PubMedBERT, and TAC 2017 Models**

Figure A. A comparison of the best-performing ClinicalBERT (CB) and PubMedBERT (PB) models with varying train/validation/test splits. PubMedBERT performs modestly better than ClinicalBERT, and more consistently across two different evaluations (our evaluation, and the TAC 2017 evaluation).

Figure B. The PubMedBERT model performs better than the models submitted for the TAC 2017 ADE Track Task as described by Demner-Fushman et al., and additionally against the compiled SIDER database.
